# Associations between GLP 1 receptor agonist therapy and surgical wound healing in a high-risk cohort

**DOI:** 10.64898/2026.04.21.26351321

**Authors:** Dominika Pullmann, Jack C. Adams, Hannah Belostotsky, Tamara Mestvirishvili, Ernest S. Chiu, Cheongeun Oh, Piul S. Rabbani

**Author notes:** contributed equally (Dominika Pullmann, Jack C. Adams). Corresponding author: Piul S. Rabbani PhD, 540 1^st^ Ave, RB-510D, New York, NY 10016.

## Abstract

**Objective:** This pilot, signal-detecting cohort study examines associations between systemic GLP-1 receptor agonist (GLP-1RA) use and surgical wound healing outcomes in high-risk surgical populations, including patients with diabetes.

**Approach:** This pilot retrospective cohort study compared adult surgery patients with non-healing postoperative wounds by their preoperative GLP-1RA use. We identified cases using standardized administrative coding and assessed wound healing outcomes through structured review of clinical notes. Outcomes included healing status, time to wound closure, and number of surgical interventions.

**Results:** The cohort included 35 non-GLP-1RA users and 16 GLP-1RA users with comparable baseline characteristics, except for significantly higher prevalence of venous insufficiency among users. Though median time to closure was similar for all patients, users required fewer surgical interventions and wound closure rates were significantly higher than in non-users. Among patients with diabetes, all GLP-1RA users healed significantly compared to non-users.

**Conclusion:** In this pilot cohort, GLP-1RA use was associated with higher unadjusted healing rates and fewer interventions, particularly among patients with diabetes. Adjusted estimates were directionally consistent but imprecise. These hypothesis-generating findings warrant prospective evaluation in larger multi-site studies before informing perioperative management.

## INTRODUCTION

Wound healing is a complex process that is critical to surgical success and patient recovery. In plastic and reconstructive surgery, successful wound closure assumes particular importance because procedures often involve extensive soft-tissue manipulation, large surface-area incisions, and tissues with compromised vascularity, such as previously scarred or irradiated fields. Patient-level comorbidities are frequent in these surgical populations and include diabetes mellitus, obesity, smoking exposure, peripheral arterial disease, venous insufficiency, chronic kidney disease, and immunosuppression. These are well-established risk factors for delayed healing, wound dehiscence, surgical site infection, and chronic non-healing wounds.^1–3^

Among these factors, type 2 diabetes mellitus (T2DM) plays a central role in impaired wound repair through mechanisms that extend beyond hyperglycemia alone. T2DM is associated with microvascular dysfunction, impaired nitric oxide bioavailability, chronic inflammation, oxidative stress, and dysregulated immune cell recruitment and polarization, all of which disrupt normal progression of wound healing through the inflammatory, proliferative, and remodeling phases.^1,3^ These interconnected metabolic and physiologic disruptions suggest why targeting glycemic control alone may be insufficient to reliably normalize wound outcomes, particularly in surgical settings.

Glucagon-like peptide-1 receptor agonists (GLP-1RAs) have become cornerstone therapies for T2DM and obesity, with rapidly expanding use across surgical populations. Beyond glucose-lowering, GLP-1RAs exert pleiotropic effects that may be directly relevant to tissue repair. Early experimental and translational studies demonstrate that GLP-1 signaling modulates inflammatory pathways, improves endothelial function, enhances angiogenesis, and promotes tissue perfusion through mechanisms involving AMP-activated protein kinase, hypoxia-inducible factor-1α, nitric oxide signaling, and cytoprotective pathways.^4–7^ In vitro studies further suggest that GLP-1RAs influence keratinocyte and fibroblast proliferation, migration, and extracellular matrix remodeling, while preclinical ischemic flap and diabetic wound models demonstrate improved tissue survival and accelerated wound closure following GLP-1RA exposure.^8–11^ Collectively, these data provide biologic plausibility for a potential role of GLP-1RAs in modifying soft-tissue healing, particularly in metabolically and vascularly compromised patients.

Despite emerging mechanistic insights and supportive preclinical evidence, limited clinical data specifically links GLP-1RA therapy to postoperative wound healing outcomes. Population-based cohort evidence has shown protective effects of GLP-1RA therapy on nonoperative wound healing outcomes in patients with T2DM including reduced rates of lower extremity amputation, diabetic foot ulcers, wound healing complications, and chronic non-healing wounds compared to alternative glucose-lowering agents or standard care.^12,13^ Other large retrospective database studies specifically examining surgical populations have reported associations between GLP-1RA use and improved surgical wound outcomes, including reduced dehiscence, readmission, and lower extremity amputation rates across heterogeneous surgical cohorts.^14–16^ In contrast, other observational analyses have demonstrated neutral or procedure-specific findings suggesting higher complication rates, including increased postoperative infections and nonunion in select orthopedic and foot-and-ankle populations, and delayed wound healing in body-contouring panniculectomy.^14,17–19^ These conflicting results underscore substantial variability in study design, patient populations, surgical context, medication exposure definitions, and wound outcome determination.

Importantly, randomized controlled trials evaluating perioperative management of GLP-1RA therapy with standardized wound healing endpoints are lacking. Much of the existing perioperative literature relies on claims or electronic health record-coded complication endpoints (e.g., infection, dehiscence, reoperation) as proxies for wound outcomes, rather than clinically adjudicated wound closure.^14,20^ Administrative claims data have well-documented limitations in sensitivity and positive predictive value for chronic conditions, particularly across repeated follow-up visits, and at discharge where 10-20% of cases have reported coding inaccuracies.^21–23^ These inaccuracies are especially consequential for defining the wound closure phenotype, which involves significant clinical heterogeneity in closure trajectory, intervention history, and care setting, limiting the ability of administrative codes to accurately capture meaningful wound healing outcomes. As a result, the impact of GLP-1RA therapy on clinically meaningful wound healing outcomes, particularly in high-risk reconstructive and soft-tissue surgery, remains poorly defined.

Given the growing prevalence of GLP-1RA use and ongoing uncertainty surrounding its relationship to surgical wound healing, additional clinical data are needed. To address this gap, we conducted a single-institution retrospective cohort study within an academic plastic surgery service to evaluate the association between GLP-1RA use and wound healing outcomes among adults with non-healing postoperative wounds requiring follow-up. We used clinically adjudicated wound closure as the primary outcome rather than claims-based complication proxies. We compared healing status at last follow-up, time to closure, and the need for additional surgical interventions between GLP-1RA users and non-users, incorporating patient-level comorbidities and concomitant diabetes therapies in adjusted analyses to better contextualize the association between GLP-1RA use and wound healing outcomes.

## CLINICAL PROBLEM ADDRESSED

This study addresses the clinical challenge of impaired healing in patients with postoperative wounds, particularly among those with comorbidities such as diabetes, in the context of GLP-1RA use and need for evidence regarding the impact of these drugs on surgical wound outcomes.

## MATERIALS AND METHODS

We conducted a single-institution retrospective cohort study within the NYU Langone Health system. The NYU Langone Institutional Review Board approved the study with a waiver of informed consent due to its retrospective nature (IRB i25-00760). This study derived data from electronic medical records and institutional data queries covering the study period from January 1, 2013 through December 31, 2024. We collected and stored study data using a secure, institutionally approved server in accordance with institutional data security policies.

The study included adult patients (≥18 years) with non-healing surgical wounds and active prescriptions for GLP-1RA or other antihyperglycemic therapies. Although not fully encompassing due to potential coding and documentation variability, we identified non-healing surgical wounds by ICD-10-CM diagnosis code T81.31XA (postoperative wound dehiscence during the initial encounter) as the most specific code to identify actively managed postoperative wound dehiscence and code T81.4XXA (postoperative infection related to a procedure, e.g., surgical site infection, during the initial encounter) to capture complementary but distinct mechanisms of non-healing surgical wounds. Index operations leading to non-healing wounds included plastic, orthopedic, vascular, oncological, neurological, and other subspecialty surgeries (**Figure 2A**). We included patients who presented to the plastic surgery service for subsequent evaluation and management (**Figure 2B**). Our criteria counted all wounds per patient, and we treated each patient as the unit of analysis. Eligibility required at least one documented follow-up visit within the plastic surgery service. The study excluded patients with incomplete medical records that precluded determination of medication exposure or wound outcome. We defined GLP-1RA exposure as an active prescription at or before the date of the index surgery and excluded postoperative initiation as a distinct exposure category.

**Figure 1.**
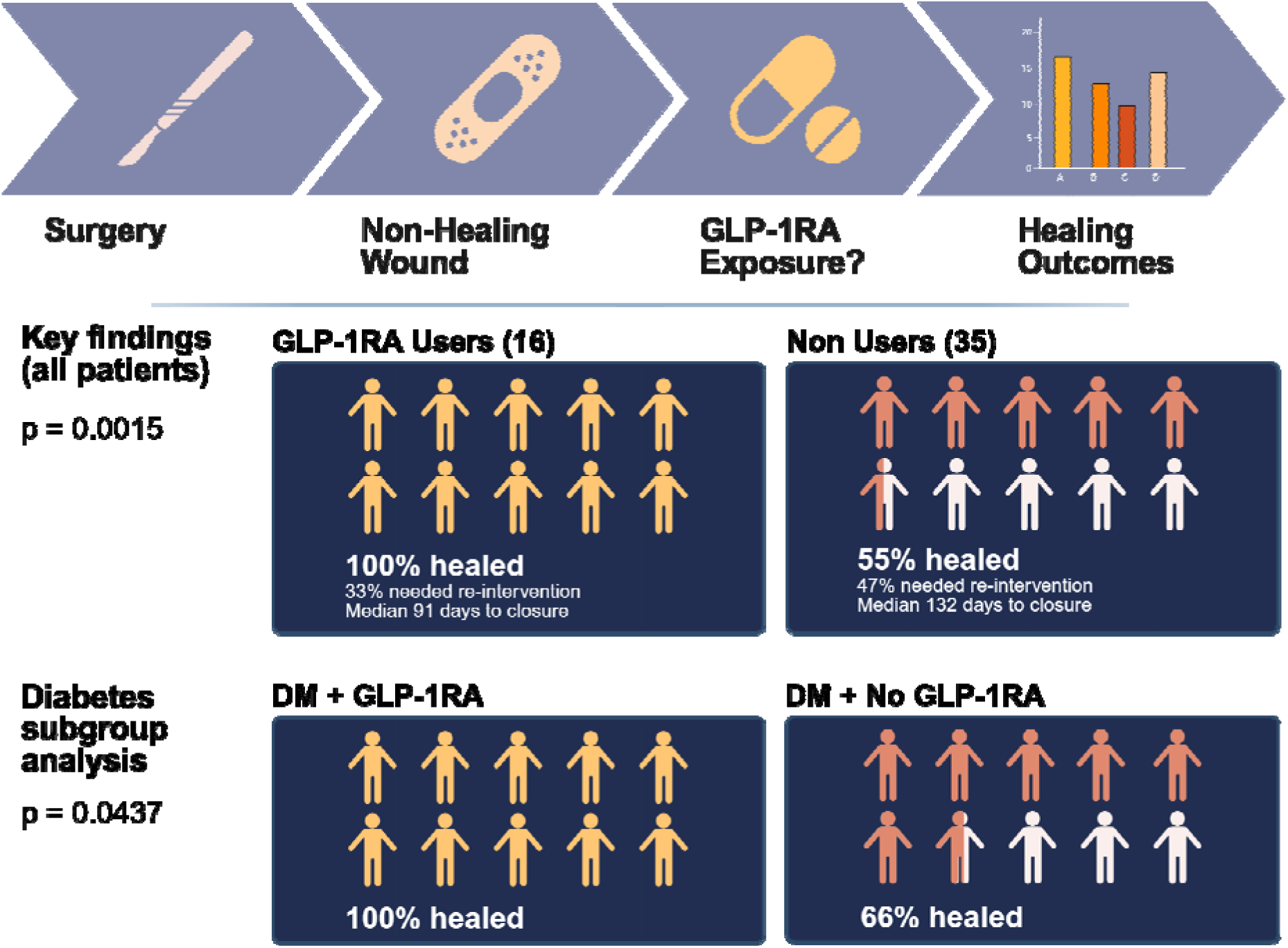
Graphical abstract. Patients taking GLP-1RA medications were more likely to have surgical wounds heal completely compared to those not taking these medications, both overall and among patients with diabetes.

**Figure 2.**
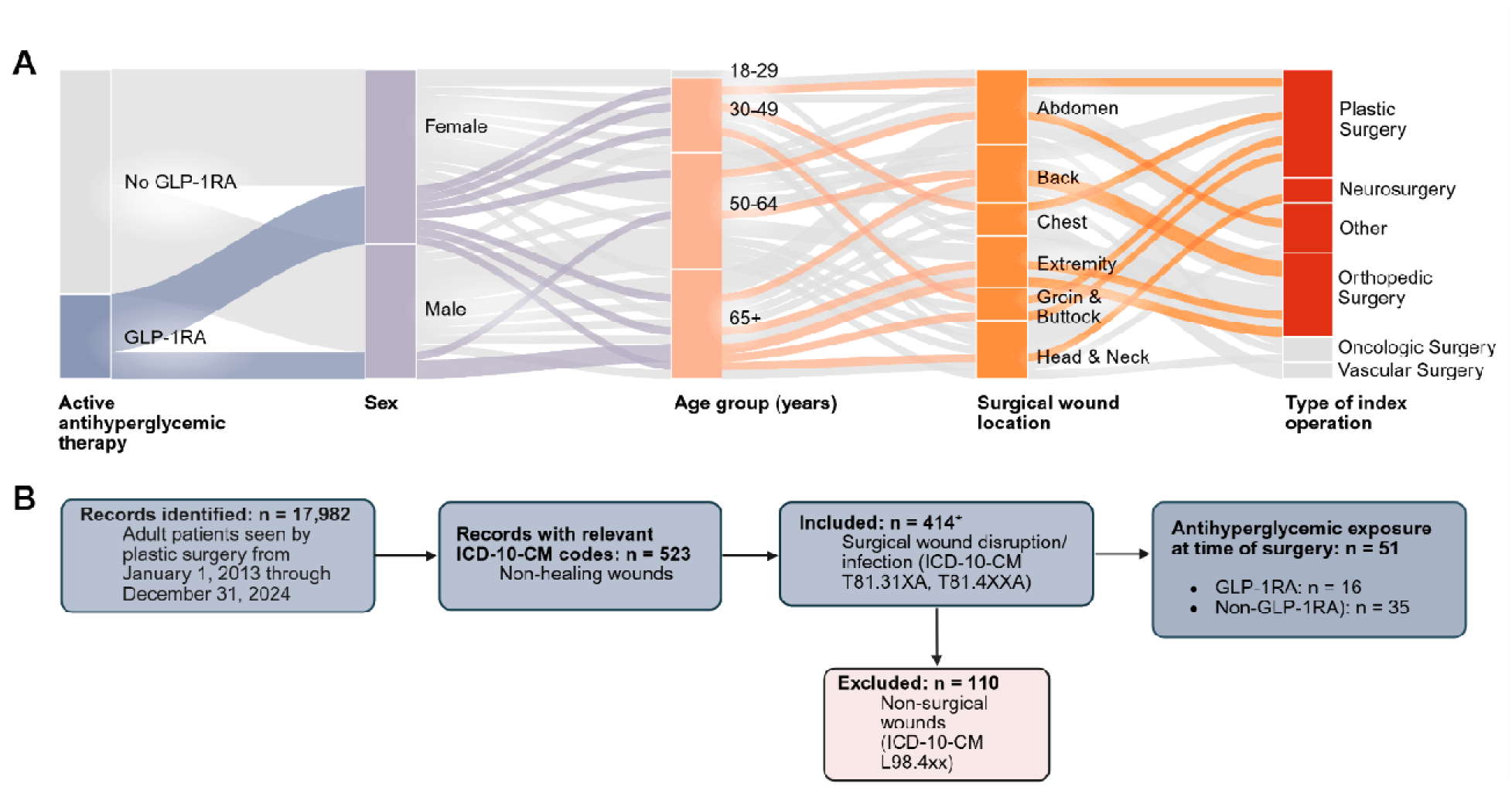
Patient selection and cohort derivation. **A**. Distribution of patients with non healing surgical wounds with active antihyperglycemic medications, the location of non-healing surgical wounds, and type of index operation. **B**. Patients seen by the plastic surgery service (2013–2024) and screened using ICD-10-CM codes. *One patient contributed two wounds to the analysis. The cohort was stratified by antihyperglycemic use at surgery: GLP-1RA users and non-users taking other antihyperglycemic therapies.

The primary outcome was wound healing status at last follow-up, which the treating plastic surgery team documented as wound closure in the electronic medical record. Secondary outcomes included time to wound closure, which was calculated from the index surgery date to the date of documented wound closure, and number of surgical interventions required for wound resolution. The study prespecified patient-level covariates based on established relevance to wound healing, including age, sex, diabetes status, body mass index, smoking status, peripheral arterial disease, venous insufficiency, chronic kidney disease, immunosuppression, wound location, and concomitant insulin and metformin use. The analysis reviewed additional contextual variables descriptively but did not include them in adjusted models due to limited sample size and data sparsity.

The analysis summarized continuous variables using means with standard deviations or medians with interquartile ranges, as appropriate, and compared groups using Mann-Whitney tests. We summarized categorical variables as frequencies and proportions and compared groups using Fisher’s exact tests. Univariable logistic regression evaluated associations between individual covariates and wound healing at last follow-up, while multivariable logistic regression assessed the association between GLP-1RA use and wound healing while adjusting for prespecified clinical covariates. Given the limited sample size and sparse data across several covariates, we interpreted adjusted estimates cautiously. The analysis defined statistical significance as a two-sided p-value <0.05. We performed statistical analyses using R (R Foundation for Statistical Computing, Vienna, Austria), including the survival package, and GraphPad Prism (version 10.6.1). Given the limited sample size, we prioritized precision-based interpretation of adjusted estimates rather than formal post-hoc power analysis, which is generally not informative for completed studies.

We conducted an exploratory propensity-score sensitivity analysis using overlap weighting to address potential residual confounding, particularly the higher prevalence of venous insufficiency among GLP-1RA users. The propensity model included age, sex, peripheral arterial disease, venous insufficiency, chronic kidney disease, insulin use, and metformin use. We assessed covariate balance using standardized mean differences before and after weighting.

## RESULTS

The study included 51 adult patients with non-healing postoperative wounds (35 non-GLP-1RA users and 16 GLP-1RA users). Among GLP-1RA users, prescribed agents included dulaglutide, semaglutide, and liraglutide; all were long-acting agents administered once-weekly or once-daily. **Table 1** summarizes baseline characteristics. Non-users and GLP-1RA users were similar in mean age (60±14 vs 56±14 years; p=0.43) and sex distribution (54% vs 63% female; p=0.58). The prevalence of key comorbidities, including diabetes, smoking status, peripheral arterial disease, chronic kidney disease, immunosuppression, malnutrition, prior radiation exposure, alcohol use disorder, and intravenous drug use, did not differ significantly between groups (all p≥0.15). Wounds involved a range of anatomic sites, most commonly the abdomen, back, and breast, with no significant difference in wound location between groups (p=0.34). Venous insufficiency was less frequent among non-users compared to GLP-1RA users (3% vs 27%; p=0.024). A non-significant trend toward lower prior surgical site infection history was observed among GLP-1RA users (21% vs 50%; p=0.07). Substantial missingness precluded meaningful comparison of baseline BMI.

**Table 1.** Baseline demographic and clinical characteristics of the study cohort stratified by GLP-1RA exposure.

| Characteristic | No GLP-1RA<br>(n=35) | GLP-1RA User<br>(n=16) | *p*-value |
| --- | --- | --- | --- |
| Age, years (mean ± SD) | 60 ± 14 | 56 ± 14 | 0.43 |
| Female sex, n (%) | 19 (54%) | 10 (63%) | 0.58 |
| **Smoking status** |  |  | **0.15** |
| Never | 20 (57%) | 8 (50%) |  |
| Former | 15 (43%) | 6 (38%) |  |
| Current | 0 (0%) | 2 (13%) |  |
| Peripheral arterial disease | 5 (14%) | 5 (31%) | 0.25 |
| Venous insufficiency | 1 (3%) | 4 (27%) | 0.024 |
| Chronic kidney disease | 10 (29%) | 5 (31%) | >0.99 |
| Immunosuppression | 0 (0%) | 0 (0%) | >0.99 |
| Malnutrition | 3 (9%) | 1 (6%) | >0.99 |
| Prior radiation exposure | 4 (11%) | 1 (6%) | >0.99 |
| Alcohol use disorder | 0 (0%) | 1 (6%) | 0.31 |
| Intravenous drug use | 0 (0%) | 0 (0%) | >0.99 |
| Prior surgical site infection | 16 (50%) | 3 (21%) | 0.07 |
| **Wound location** |  |  | **0.34** |
| Abdomen | 9 (26%) | 4 (25%) |  |
| Back | 5 (14%) | 0 (0%) |  |
| Breast (L/R) | 3 (9%) | 2 (13%) |  |
| Neck | 2 (6%) | 0 (0%) |  |
| Lower extremity | 2 (6%) | 3 (19%) |  |
| Other | 13 (37%) | 7 (44%) |  |
| Continuous variables are reported as mean ± standard deviation and compared using Wilcoxon rank-sum tests. Categorical variables are reported as n (%) and compared using Fisher's exact tests. Wound location categories include abdomen, back, breast (left/right), neck, lower extremity, and other. Bolded *p*-values denote overall comparisons for multi-level categorical variables. |  |  |  |

In unadjusted comparison, median time to wound closure did not differ significantly between all non-users and GLP-1RA users (132 vs 91 days; p=0.05; **Figure 3A**). GLP-1RA users achieved higher rates of wound healing at follow-up (100% vs 54.3%; absolute risk difference 45.7%; number needed to treat 2.2; p=0.0015) and had fewer wounds requiring surgical interventions to achieve wound resolution (33% vs 47%; absolute risk difference 14.0%; p=0.37; **Figures 3B-C; Table S1**). Given the retrospective and observational design, the number needed to treat is presented as a descriptive effect-size measure rather than a causal estimate. Among patients with diabetes, GLP-1RA users demonstrated higher healing rates than diabetic non-users (100% vs 66%; absolute risk difference approximately 34%; number needed to treat approximately 3; p=0.0437; **Figure 3D**).

**Figure 3.**
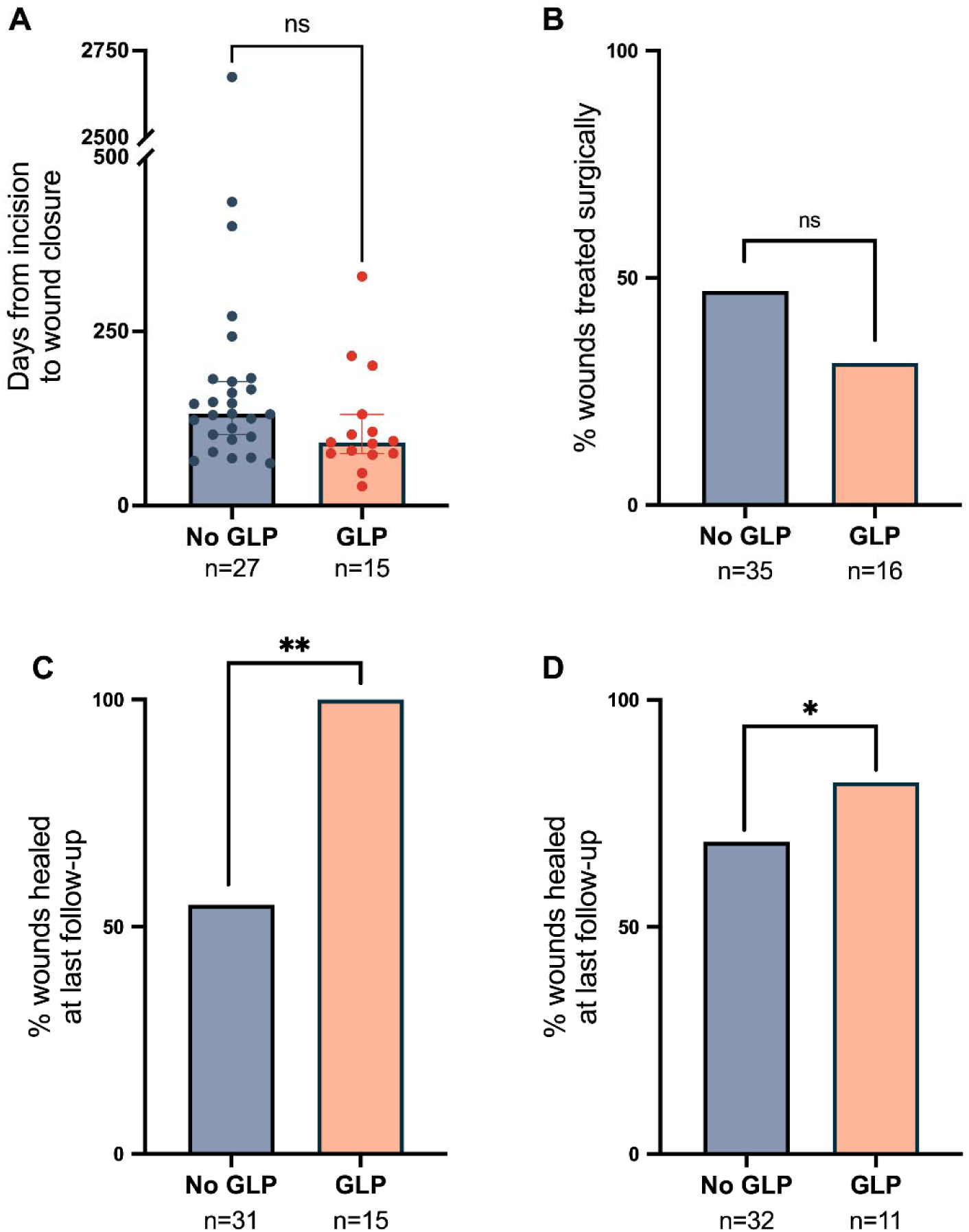
Wound healing outcomes in patients with and without diabetes, stratified by GLP-1RA exposure. **A**. Time from incision to wound closure (days) among non-diabetic patients, displayed as individual data points with mean ± SEM. **B**. Proportion of wounds requiring surgical intervention among non-diabetic patients. **C**. Percentage of wounds healed at last follow-up among non-diabetic patients, *p* < 0.01). **D**. Percentage of wounds healed at last follow-up among diabetic patients, *p* < 0.05. Statistical comparisons by Fisher’s exact test (B–D) and Wilcoxon rank-sum test (A).

Univariable logistic regression analyses evaluating factors associated with wound healing are shown in **Figure 4A**. GLP-1RA use was associated with higher odds of wound healing, although this association did not reach statistical significance (OR 2.80; 95% CI 0.62–19.92; p=0.22) (**Figure 4B**). Age, sex, smoking status, peripheral arterial disease, venous insufficiency, chronic kidney disease, insulin use, and metformin use were not significantly associated with healing. Several estimates demonstrated wide confidence intervals, reflecting limited sample size and sparse data across predictor strata. Smoking status, in particular, contained sparse data with zero or very small cell counts in one or more categories (**Table 1**), producing a non-estimable odds ratio for current smokers; this finding should be interpreted with caution.

**Figure 4.**
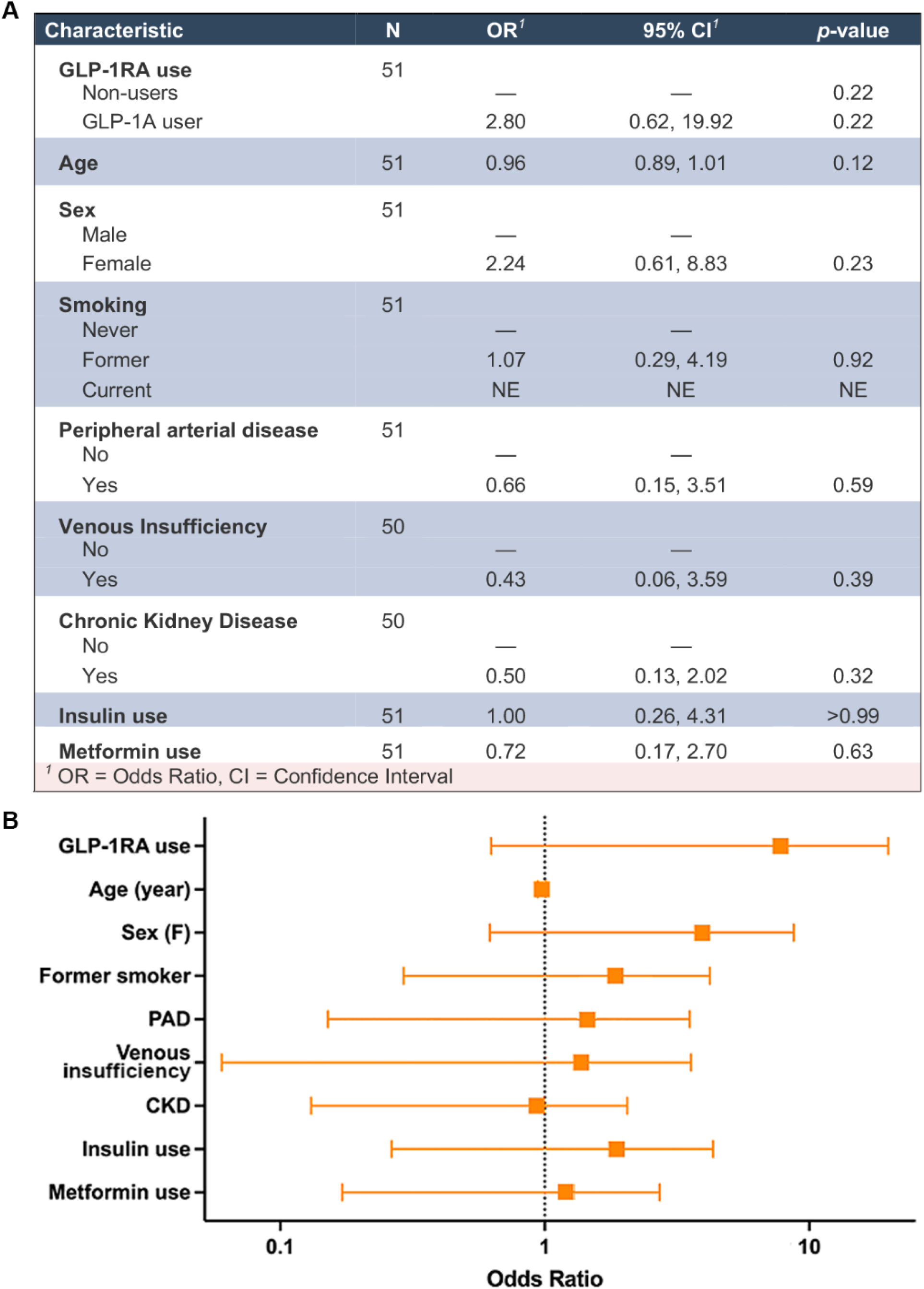
Univariable logistic regression analysis of factors associated with wound healing at last follow-up. **A**. This table presents OR with 95% CI and *p*-values for each predictor. For categorical variables, “—” denotes reference. The current smoker category contained sparse data with zero or very small cell counts (Table 1), producing a non-estimable odds ratio (NE); these findings should be interpreted with caution. The adjusted model and forest plot N excluded this variable. N varies by predictor due to missing data. **B**. Forest plot of odds ratios on a logarithmic scale. Point estimates represent OR; horizontal bars represent 95% CI. The dotted vertical line at OR = 1.0 represents the null hypothesis of no association.

Adjusted logistic regression analyses are summarized in **Table 2** and evaluated age, sex, diabetes, BMI, smoking status, peripheral arterial disease, venous insufficiency, chronic kidney disease, immunosuppression, wound location, and concomitant insulin and metformin use. GLP-1RA use corresponded to higher odds of wound healing, although this association did not reach statistical significance (OR 11.6; 95% CI 0.54–593.8; p=0.15). Estimates for multiple covariates demonstrated wide confidence intervals, reflecting limited sample size and sparse data within several predictor strata. In particular, diabetes status exhibited evidence of model instability consistent with complete or quasi-complete separation, which limited reliable estimation of its independent effect. Venous insufficiency showed a strong inverse point estimate for healing but did not reach statistical significance in adjusted analyses (OR 0.03; p=0.07). Limited sample size, missing data, and lack of variation across several predictors prevented reliable estimation of expanded sensitivity models incorporating additional secondary and contextual variables. Accordingly, we interpreted adjusted findings cautiously and presented these as exploratory. The wide confidence interval for GLP-1RA use in the adjusted model (OR 11.6; 95% CI 0.54–593.8) reflects imprecision driven by limited sample size and sparse covariate data rather than evidence against an association; the point estimate remained large and directionally consistent with unadjusted findings throughout.

**Table 2.** Adjusted odds of healed wounds at last follow-up (logistic regression).

| Variable | Adjusted OR | 95% CI<sup>1</sup> | p-value |
| --- | --- | --- | --- |
| GLP-1RA use | 11.6 | 0.54–593.8 | 0.15 |
| Age (per year) | 0.98 | 0.87–1.09 | 0.72 |
| Female sex | 0.61 | 0.03–11.46 | 0.73 |
| Smoking (former vs never) | 0.42 | 0.03–3.88 | 0.46 |
| Peripheral arterial disease | 0.46 | 0.04–4.25 | 0.50 |
| Venous insufficiency | 0.03 | 0.00–0.96 | 0.07 |
| Chronic kidney disease | 0.48 | 0.05–4.44 | 0.52 |
| Insulin use | 1.31 | 0.07–29.25 | 0.86 |
| Metformin use | 0.86 | 0.03–26.02 | 0.93 |

<sup>1</sup> Wide confidence intervals reflect limited sample size and sparse data across predictor strata.

In an exploratory overlap-weighted propensity-score sensitivity analysis, covariate balance improved substantially after weighting, with all standardized mean differences reduced below 0.1 (**Table S2**). GLP-1RA use remained positively associated with wound healing in the weighted analysis (OR 6.99; 95% CI 0.59–82.4; p=0.12; **Table S3**), consistent in direction with primary analyses. However, overlap weighting substantially reduced the effective sample size (treated approximately 6.3; control approximately 12.0), reflecting limited covariate overlap in this small cohort, and we interpreted these results as exploratory and hypothesis-generating.

## DISCUSSION

In this single-institution retrospective cohort study of patients with non-healing postoperative wounds managed within a plastic surgery service, systemic GLP-1RA use was associated with higher unadjusted healing rates and fewer subsequent surgical interventions, particularly among patients with diabetes. However, after adjusting for patient-level comorbidities and concomitant diabetes therapies, the association between GLP-1RA use and wound healing did not reach statistical significance, with wide confidence intervals reflecting limited sample size and sparse data. Taken together, these findings suggest a potential beneficial signal that warrants cautious interpretation and further study rather than definitive conclusions.

The observed trends toward improved healing among GLP-1RA users are biologically plausible and align with emerging mechanistic data. Physiologic wound healing requires tightly coordinated immune modulation and inflammatory resolution, angiogenesis, and extracellular matrix remodeling. Disruption of these processes, particularly in patients with diabetes, vascular disease, or chronic inflammation, is associated with impaired healing and chronic wound formation.^1–4,7,10^ GLP-1RAs may address multiple pathologies characteristic of diabetic chronic ulcers, including oxidative stress, chronic inflammation, neuropathy, and endothelial dysfunction.^7–11^

The limited number of preclinical studies to date support this multifactorial impact. Experimental diabetic wound models demonstrated accelerated wound closure and improved angiogenesis, with both in vivo and in vitro studies showing improved ischemic tissue flap survival and enhanced keratinocyte migration and proliferation after GLP-1RA exposure.^7–9^ A recent narrative and systematic review further describes cellular and molecular signaling pathways linking systemic GLP-1RAs to wound healing mechanisms in diabetic ulcers.^25^ Together with the existing literature, these data suggest that GLP-1RAs may influence wound healing through mechanisms independent of weight loss or glucose lowering alone, providing a biologic framework for the improved healing rates observed in our cohort.

Clinical evidence evaluating GLP-1RA use and postoperative wound outcomes remains limited and heterogeneous. Comprehensive population-based observational studies have reported associations between GLP-1RA therapy and improved postoperative outcomes, including reduced wound dehiscence and lower readmission rates across broad surgical populations.^14,15^ More recently, large-scale data have also suggested potential limb-protective effects of GLP-1RAs in high-risk diabetic populations.^13,26,27^ In a large retrospective cohort study, GLP-1RA use was associated with lower risks of diabetic foot ulcers, lower-extremity amputations and mortality, compared with sodium-glucose transport 2 inhibitor therapy, particularly among patients with peripheral artery disease or preexisting ulcers.^12^ In contrast, other procedure-specific cohorts have demonstrated neutral or conflicting findings, including increased infection rates following certain orthopedic procedures and delayed wound healing in body-contouring surgeries.^17,18^ These discrepancies likely reflect differences in surgical context, baseline patient risk, exposure and outcome definitions rather than true biologic inconsistency.

Our findings add to this evolving literature by focusing specifically on an actively managed non-healing surgical wound cohort, a clinically and biologically distinct phenotype characterized by persistent wound failure despite ongoing intervention, and one that is underrepresented in prior GLP-1RA surgical outcome studies (**Table S4**). Defining the actively managed non-healing surgical wound cohort as a target population for future GLP-1RA research may yield more biologically interpretable findings, than analyses that use surgical procedure type as the unit of analysis without accounting for wound healing trajectory.

Clinical reports thus far regarding surgical outcomes in patients on GLP-1RA therapy largely derive from orthopedic, spine, or broad surgical registries.^14,19,20,28,29^ While large-cohort studies offer valuable representative power, wound healing outcomes in these datasets are approximated through administrative coding rather than clinical adjudication. Administrative classification structures do not fully capture the granularity of individual clinical trajectories and are a recognized methodological limitation of administrative health data.^22,23^ Importantly, unlike prior studies that analyzed outcomes at the wound level, we conducted our analysis at the patient level, addressing a key methodological challenge in wound healing research where multiple wounds per patient can introduce non-independence and analytic bias. The absence of statistically significant adjusted associations in our study should therefore be interpreted in light of limited power rather than as evidence against a biologic effect.

From a clinical perspective, perioperative GLP-1RA management has largely centered on aspiration risk related to delayed gastric emptying rather than wound outcomes.^30^ Earlier consensus recommendations favored temporary cessation before elective procedures, whereas more recent multi-society guidance supports continuation for most patients with individualized risk modifying strategies (e.g., a 24-hour liquid diet, anesthesia plan adjustments, or procedure delay in select high-risk patients).^14,31^ Our study did not evaluate aspiration or anesthesia-related outcomes and therefore should not be used to inform anesthesia-focused perioperative guidance. However, in this cohort, preoperative GLP-1RA use was not associated with impaired wound healing and showed a positive association with healing in select populations, particularly patients with diabetes (**Figure S1**). These findings suggest that for high-risk patients with impaired healing biology, who may develop non-healing wounds even after low-risk procedures, pre-operative GLP-1RA discontinuation may not be warranted based on wound healing concerns alone. As continuation may be associated with favorable healing trajectories, further investigation is necessary before recommending changes to existing perioperative protocols.

While preoperative decision-making has appropriately prioritized aspiration risk in the immediate perioperative period, the postoperative period presents distinct challenges to patient recovery. Surgical wound healing extends beyond the acute perioperative window, progressing over weeks to months and years. This temporal distinction warrants reconsideration of the optimal timing for GLP-1RA reinitiation. For patients whose GLP-1RAs therapy was discontinued preoperatively, earlier postoperative resumption may support more favorable wound healing trajectories, particularly for those with diabetes or other comorbidities that impair tissue repair.^7–11^ Patients not previously on GLP-1RAs may show more favorable healing trajectories with postoperative initiation, particularly those with elevated wound complication risks such as people who use tobacco or with inadequate glycemic control on alternative regimens. Incorporating wound healing considerations into perioperative decision-making could transform GLP-1RA management into a dynamic strategy addressing both immediate safety and downstream tissue repair concerns, potentially improving surgical outcomes.

This study has several important limitations. The cohort size was small, and multiple covariates showed sparse data or limited variability, yielding wide confidence intervals and evidence of model instability. Quasi-complete separation particularly affected diabetes status and venous insufficiency, limiting reliable estimation of independent effects. Attempts to expand the cohort to improve statistical stability identified no additional eligible cases meeting all inclusion criteria within the study timeline. This analysis therefore represents a pilot, signal-detecting study explicitly, with confirmatory work planned in larger institutional and multi-site cohorts. We could not further characterize exposure timing relative to wound evolution, or dose or duration of use within the preoperative exposure window. Additionally, some non-users may have initiated GLP-1RA therapy during their postoperative course, representing crossover contamination that would misclassify exposed patients as unexposed and underestimate true between-group differences in healing rates. Although all included GLP-1RA users received long-acting agents, we could not assess differential effects by agent type or dose within this limited sample. GLP-1RA users, as we defined, also differed from non-users in baseline characteristics, most notably a higher prevalence of venous insufficiency, which may have attenuated adjusted effect estimates despite favorable unadjusted outcomes. Beyond these measured confounders, GLP-1RA users may differ from non-users in ways that favor wound healing that were not captured in our data, including greater health system engagement, higher medication adherence, and GLP-1RA-associated weight loss. These features highlight the difficulty of disentangling medication effects from underlying patient complexity in retrospective wound-care datasets and reinforce the need for larger cohorts with sufficient power to support robust multivariable modeling. Importantly, adjusted effect estimates for GLP-1RA use remained directionally consistent with unadjusted findings, suggesting statistical imprecision rather than absence of a biologic association. Rather than performing post-hoc power calculations, which are generally not informative for completed analyses, we emphasize a precision-based interpretation of the adjusted model. Additional limitations include the retrospective design, single-institution single-service setting, incomplete data for certain variables (e.g., BMI), inability to fully characterize medication exposure timing, duration, and adherence, and limited wound-level granularity based on available documentation. These limitations preclude causal inference and underscore the exploratory nature of our findings.

To acquire the evidence needed to inform patient counseling and surgical planning with more predictable outcomes, future research needs to prioritize larger, prospective, multicenter cohorts with standardized wound healing endpoints and analytic strategies accounting for clustering of wounds within individual patients. Studies should characterize preoperative exposure duration, agent class, dose, and timing of postoperative reinitiation as exposure dimensions as these are most likely to determine whether and when GLP-1RA therapy meaningfully modifies the wound healing trajectory. The improved unadjusted healing rates observed in our cohort, combined with other retrospective studies, highlight the need for translational research to establish causation and mechanistic insights downstream of systemic GLP-1RA exposure. As a pilot, signal-detecting cohort, our findings provide hypothesis-generating evidence to support further investigation into the role of GLP-1RAs as potential modifiers of surgical wound healing.

## Data Availability

Data produced in the present study are not available upon request to the authors.

## ACKNOWLEDGMENTS

The authors thank Thomas Calahan for facilitating IRB review and approval for this study, and Vincent Setang for assisting with retrieval of patient records.

## AUTHOR CONTRIBUTIONS

Conceptualization, project administration, resources, supervision, funding acquisition: PSR

Conceptualization, data curation, methodology, investigation, analysis, visualization: JCA, DP

Data curation, methodology, investigation, analysis, validation, visualization: JCA, DP, HB, TM, CO, PSR

Conceptualization, supervision: EC

Writing, original draft: DP, JCA, CO, PSR

Writing, review and editing: DP, JCA, HB, TM, EC, CO, PSR

## ABBREVIATIONS AND ACRONYMS

AMP: adenosine monophosphate
BMI: body mass index
CI: confidence interval
EMR: electronic medical record
GLP-1: glucagon-like peptide-1
GLP-1RA: glucagon-like peptide-1 receptor agonist
HIF-1α: hypoxia-inducible factor-1α
ICD-10-CM: International Classification of Diseases, Tenth Revision, Clinical Modification
IRB: Institutional Review Board
OR: odds ratio
SMD: standard mean difference
T2DM: type 2 diabetes mellitus

## REFERENCES

1. Janis JE, Harrison B. Wound Healing: Part I. Basic Science. Plastic & Reconstructive Surgery. 2016;138(3S):9S–17S. doi:10.1097/PRS.0000000000002773

2. Basak M, Sharma A, Laskar MA. Recent Advances in Wound Healing and Wound Care: A Comprehensive Review of Mechanisms and Therapeutic Innovations. CIS. 2025;03:e2210299X392533. doi:10.2174/012210299X392533250818055951

3. Kibibi ML. Exploration of Management Strategies for Type 2 Diabetes among Wounded Patients: A Review. IDOSR-JAS. 2024;9(3):48–52. doi:10.59298/IDOSRJAS/2024/9.3.4852000

4. Gong B, Li C, Shi Z, et al. GLP-1 receptor agonists: exploration of transformation from metabolic regulation to multi-organ therapy. Front Pharmacol. 2025;16:1675552. doi:10.3389/fphar.2025.1675552

5. Park B, Krishnaraj A, Teoh H, et al. GLP-1RA therapy increases circulating vascular regenerative cell content in people living with type 2 diabetes. American Journal of Physiology-Heart and Circulatory Physiology. 2024;327(2):H370–H376. doi:10.1152/ajpheart.00257.2024

6. Cedirian S, Donati M, Rapparini L, et al. Benefit-Risk Assessment of GLP-1 Receptor Agonists: Implications for Dermatologists and Plastic Surgeons. Dermatol Ther (Heidelb). 2025;15(11):3173–3193. doi:10.1007/s13555-025-01537-5

7. Huang H, Wang L, Qian F, et al. Liraglutide via Activation of AMP-Activated Protein Kinase-Hypoxia Inducible Factor-1α-Heme Oxygenase-1 Signaling Promotes Wound Healing by Preventing Endothelial Dysfunction in Diabetic Mice. Front Physiol. 2021;12:660263. doi:10.3389/fphys.2021.660263

8. Zhu X, Hu X, Lou J, et al. Liraglutide, a TFEB-Mediated Autophagy Agonist, Promotes the Viability of Random-Pattern Skin Flaps. Moon KM, ed. Oxidative Medicine and Cellular Longevity. 2021;2021(1):6610603. doi:10.1155/2021/6610603

9. Zhang Q, Zhang C, Kang C, et al. Liraglutide Promotes Diabetic Wound Healing via Myo1c/Dock5. Advanced Science. 2024;11(39):2405987. doi:10.1002/advs.202405987

10. Yu M, Huang J, Zhu T, et al. Liraglutide-loaded PLGA/gelatin electrospun nanofibrous mats promote angiogenesis to accelerate diabetic wound healing *via* the modulation of miR-29b-3p. Biomater Sci. 2020;8(15):4225–4238. doi:10.1039/D0BM00442A

11. Liu T, Zhou L, Chen Y, Lin J, Zhu H. Semaglutide outperforms insulin in restoring neutrophil function against implant-related infection in diabetic and obese mice: experimental research. International Journal of Surgery. 2025;111(1):273–282. doi:10.1097/JS9.0000000000001896

12. Hong AT, Luu IY, Lin F, et al. Differential Effect of GLP-1 Receptor Agonists and SGLT2 Inhibitors on Lower-Extremity Amputation Outcomes in Type 2 Diabetes: A Nationwide Retrospective Cohort Study. Diabetes Care. 2025;48(10):1728–1736. doi:10.2337/dc25-0292

13. Lewis JE, Omenge DK, Patterson AR, et al. The impact of semaglutide on wound healing in diabetes related foot ulcer patients: A TriNetX database study. Diab Vasc Dis Res. 2025;22(2):14791641251322909. doi:10.1177/14791641251322909

14. Aschen SZ, Zhang A, O’Connell GM, et al. Association of Perioperative Glucagon-like Peptide-1 Receptor Agonist Use and Postoperative Outcomes. Annals of Surgery. 2025;281(4):600–607. doi:10.1097/SLA.0000000000006614

15. White CJK, Choudhury A, Bollepalli H, Kodra JD, Burton AT, Kraus JC. Preoperative GLP-1 Receptor Agonist Use and Postoperative Outcomes Following Operative Ankle Fracture Repair in Patients With Type 2 Diabetes: A National Database Study. Foot Ankle Int. 2025;46(11):1265–1274. doi:10.1177/10711007251364187

16. Hiredesai AN, Howlett CP, Kisiel S, et al. Glucagon-like Peptide-1 Receptor Agonist Therapy Is Associated With Fewer Medical Complications After Carpal Tunnel Release in Diabetic Patients. Hand (New York, N,Y). Published online July 16, 2025:15589447251350181. doi:10.1177/15589447251350181

17. Levidy M, Vatsia S, Tucker S, Rowe N, Aynardi M, MacDonald A. The Impact of GLP-1 Agonist Therapy in Tibiotalar and Subtalar Fusion: A Propensity-Matched Analysis. Foot & Ankle Orthopaedics. 2024;9(4):2473011424S00536. doi:10.1177/2473011424S00536

18. Koenig ZA, Rashid S, Hobbs GR, Uygur HS. Perioperative GLP-1 Receptor Agonist Use & Surgical Outcomes in Non-bariatric Abdominal Panniculectomy: A 10-Year Retrospective Analysis. Plastic & Reconstructive Surgery. Published online August 26, 2025. doi:10.1097/PRS.0000000000012405

19. Pereira DE, Tummala S, Mittal MM, et al. Perioperative glucagon-like Peptide-1 receptor agonist use and clinical outcomes following lower extremity fracture fixation: A large retrospective cohort study with two year follow up. Injury. 2025;56(11):112746. doi:10.1016/j.injury.2025.112746

20. Rashid Z, Woldesenbet S, Khalil M, et al. Impact of Preoperative Glucagon-Like Peptide-1 Receptor Agonist on Outcomes Following Major Surgery. World j surg. 2025;49(3):698–707. doi:10.1002/wjs.12484

21. Wei WQ, Teixeira PL, Mo H, Cronin RM, Warner JL, Denny JC. Combining billing codes, clinical notes, and medications from electronic health records provides superior phenotyping performance. J Am Med Inform Assoc. 2016;23(e1):e20–e27. doi:10.1093/jamia/ocv130

22. Kuang A, Xu C, Southern DA, Sandhu N, Quan H. Validated administrative data based ICD-10 algorithms for chronic conditions: A systematic review. J Epidemiol Popul Health. 2024;72(4):202744. doi:10.1016/j.jeph.2024.202744

23. Wang Y, Song Y, Siu R, et al. Validation of 13 102 International Classification of Diseases, Tenth Revision, Clinical Modification codes using a large language model-based system. J Am Med Inform Assoc. 2026;33(5):947–956. doi:10.1093/jamia/ocag008

24. Adams JC, Pullmann D, Belostotsky H, et al. Effects of Glucagon-Like Peptide-1 (GLP-1) Agonists on Surgical Wound Healing: A Single Institution Pilot Study. medRxiv. Published online January 1, 2026:2026.04.21.26351321. doi:10.64898/2026.04.21.26351321

25. Gruzmark FS, Beraja GE, Jozic I, Lev-Tov HA. Exploring the Role of GLP-1 Agents in Managing Diabetic Foot Ulcers: A Narrative and Systematic Review. Wound Repair Regen. 2025;33(5):e70085. doi:10.1111/wrr.70085

26. Werkman NCC, Driessen JHM, Stehouwer CDA, et al. The use of sodium-glucose co-transporter-2 inhibitors or glucagon-like peptide-1 receptor agonists versus sulfonylureas and the risk of lower limb amputations: a nation-wide cohort study. Cardiovasc Diabetol. 2023;22(1):160. doi:10.1186/s12933-023-01897-2

27. Dhatariya K, Bain SC, Buse JB, et al. The Impact of Liraglutide on Diabetes-Related Foot Ulceration and Associated Complications in Patients With Type 2 Diabetes at High Risk for Cardiovascular Events: Results From the LEADER Trial. Diabetes Care. 2018;41(10):2229–2235. doi:10.2337/dc18-1094

28. Kishan A, Khela HS, Carayannopoulos NL, et al. Association of Glucagon-like Peptide-1 Receptor Agonist Use with Complications Following Thoracic and/or Lumbar Spinal Fusion for Degenerative Spine Disease: A BMI-Stratified Retrospective Study. Spine. Published online September 4, 2025. doi:10.1097/BRS.0000000000005494

29. Wiener JM, Sanghvi PA, Vlastaris K, et al. Glucagon-Like Peptide-1 Receptor Agonist Medications Alter Outcomes of Spine Surgery: A Study Among Over 15,000 Patients. Spine. 2025;50(13):871–880. doi:10.1097/BRS.0000000000005283

30. Klein SR, Hobai IA. Semaglutide, delayed gastric emptying, and intraoperative pulmonary aspiration: a case report. Can J Anaesth. 2023;70(8):1394–1396. doi:10.1007/s12630-023-02440-3

31. Kindel TL, Wang AY, Wadhwa A, et al. Multi-society clinical practice guidance for the safe use of glucagon-like peptide-1 receptor agonists in the perioperative period. Surg Endosc. 2025;39(1):180–183. doi:10.1007/s00464-024-11263-2

32. Aschen SZ, Zhang A, O’Connell GM, et al. Association of Perioperative Glucagon-like Peptide-1 Receptor Agonist Use and Postoperative Outcomes. Annals of Surgery. 2025;281(4):600–607. doi:10.1097/SLA.0000000000006614

33. White CJK, Choudhury A, Bollepalli H, Kodra JD, Burton AT, Kraus JC. Preoperative GLP-1 Receptor Agonist Use and Postoperative Outcomes Following Operative Ankle Fracture Repair in Patients With Type 2 Diabetes: A National Database Study. Foot Ankle Int. 2025;46(11):1265–1274. doi:10.1177/10711007251364187

34. Hiredesai AN, Howlett CP, Kisiel S, et al. Glucagon-like Peptide-1 Receptor Agonist Therapy Is Associated With Fewer Medical Complications After Carpal Tunnel Release in Diabetic Patients. Hand (New York, N,Y). Published online July 16, 2025:15589447251350181. doi:10.1177/15589447251350181

35. Levidy M, Vatsia S, Tucker S, Rowe N, Aynardi M, MacDonald A. The Impact of GLP-1 Agonist Therapy in Tibiotalar and Subtalar Fusion: A Propensity-Matched Analysis. Foot & Ankle Orthopaedics. 2024;9(4):2473011424S00536. doi:10.1177/2473011424S00536

36. Koenig ZA, Rashid S, Hobbs GR, Uygur HS. Perioperative GLP-1 Receptor Agonist Use & Surgical Outcomes in Non-bariatric Abdominal Panniculectomy: A 10-Year Retrospective Analysis. Plastic & Reconstructive Surgery. Published online August 26, 2025. doi:10.1097/PRS.0000000000012405

37. Rashid Z, Woldesenbet S, Khalil M, et al. Impact of Preoperative Glucagon-Like Peptide-1 Receptor Agonist on Outcomes Following Major Surgery. World j surg. 2025;49(3):698–707. doi:10.1002/wjs.12484

38. Wiener JM, Sanghvi PA, Vlastaris K, et al. Glucagon-Like Peptide-1 Receptor Agonist Medications Alter Outcomes of Spine Surgery: A Study Among Over 15,000 Patients. Spine. 2025;50(13):871–880. doi:10.1097/BRS.0000000000005283

