## Supplemental Materials for "Associations between GLP 1 receptor agonist therapy and surgical wound healing in a high-risk cohort"

| **Group** | **N** | **Healed (n)** | **Healing Proportion** |
| --- | --- | --- | --- |
| No GLP-1RA | 35 | 25 | 0.714 |
| GLP-1RA user | 16 | 14 | 0.875 |

**Table S1. Unadjusted healing outcomes by GLP-1RA exposure.**

| **Variable** | **Unadjusted SMD** | **Adjusted SMD*^1^*** |
| --- | --- | --- |
| Propensity score | 2.056 | 0.053 |
| Age | -0.535 | 0.000 |
| Female sex | 0.041 | 0.000 |
| Peripheral arterial disease | 0.120 | 0.000 |
| Venous insufficiency | 0.237 | 0.000 |
| Chronic kidney disease | -0.028 | 0.000 |
| Insulin use | -0.153 | 0.000 |
| Metformin use | -0.528 | 0.000 |
| ***^1^*** SMD; standard mean differences |  |  |

**Table S2. Covariate balance before and after overlap weighting.**

| **Variable** | **OR** | **95% CI** | ***p*-value** |
| --- | --- | --- | --- |
| GLP-1RA use | 6.99 | 0.59–82.4 | 0.12 |

**Table S3. Propensity-weighted logistic regression.**

| **Study** | **Setting** | **Population** | **Sample size** | **Data source** | **Exposure definition** | **Outcome definition** | **Unit** | **Analytic approach** | **Primary finding** |
| --- | --- | --- | --- | --- | --- | --- | --- | --- | --- |
| **Present study** | **Non-healing postoperative wounds** | **Established non-healing wounds across diverse index operations** | **16 GLP-1RA, 35 nonusers** | **Single-institution chart review** | **Active prescription at or before index surgery** | **Clinically adjudicated wound closure documented by treating plastic surgery team** | **Patient** | **Logistic regression; exploratory overlap weighting** | **Favorable (unadjusted). ↑ healing (100% vs 54.3%; NNT 2.2). Adjusted estimates imprecise.** |
| Aschen 2025 | Mixed surgical | T1DM or T2DM | 35,020 procedures after 1:1 PSM (17,510 GLP-1RA) | Two-institution EHR | Active prescription with end date after surgery | ICD-10 codes within 180 days (dehiscence, infection, hematoma, bleeding); 30-day readmission | Procedure | 1:1 PSM; GEE | Favorable. ↓ readmission (RR 0.88), ↓ dehiscence (RR 0.71), ↓ hematoma (RR 0.44). |
| White 2025 | Ankle fracture | T2DM | 1,107 vs 1,107 after 1:1 PSM | TriNetX (multi-institution EHR) | Prescription within 180 days pre-op | ICD-10 codes (0–90 d medical; 90 d–6 mo operative) | Patient | 1:1 PSM; FDR correction | Mixed. ↓ mortality (OR 0.33), ↑ falls (OR 1.32). No difference in infection or wound healing. |
| Hiredesai 2025 | Carpal tunnel release | T2DM with CTS | 25,229 vs 25,229 after exact matching | PearlDiver (national claims) | Any prescription during study period | Composite ICD-coded 90-day medical and surgical complications; 2-year revision | Patient | Exact matching; logistic regression | Favorable. ↓ 90-day medical complications (OR 0.90), ↓ readmissions (OR 0.85). |
| Levidy 2024 (abstract) | Tibiotalar/subtalar fusion | Diabetic | 708 vs 708 after PSM | TriNetX Diamond | Not specified | ICD codes within 1 year: infection (T81.4), pseudoarthrosis (M96.0) | Patient | PSM; chi-squared | Unfavorable for infection. ↑ infection (9.0% vs 5.6%). No difference in pseudoarthrosis. |
| Koenig 2025 | Nonbariatric panniculectomy | Excluded bariatric and hernia | 81 GLP-1RA, 292 nonusers (no PSM) | Single-institution chart review | Continuous use >30 days pre-op | Chart-based clinically adjudicated wound outcomes (delayed healing, dehiscence, seroma, SSI, hematoma) | Patient | Multivariable logistic regression | Mixed. ↑ delayed healing (OR 2.79), ↓ seroma (OR 0.32). |
| Rashid 2025 | Major surgery (CABG, AAA, colectomy, pancreatectomy, pneumonectomy) | Age <65 | 2,943 vs 5,863 after PSM | IBM MarketScan (commercial claims) | Claim 1 yr to 15 d pre-op | ICD-coded 30-day complications; 30-day readmission | Patient | PSM; logistic regression | Neutral. No association with overall complications (OR 0.99). No difference in readmissions. |
| Wiener 2025 | Thoracolumbar spinal fusion | T2DM on metformin, stratified by obesity | Obese 1,560 vs 1,560; non-obese 703 vs 703 | TriNetX | Prescription at index plus post-index refill | ICD-coded 1-year outcomes (infection, readmission, revision, fracture, pain, mobility) | Patient | 1:1 PSM; Cox proportional hazards | Favorable. ↓ infection, ↓ revisions, ↓ readmissions, ↓ mobility issues. |
| Hong 2025 | Lower-extremity outcomes | T2DM (comparator: SGLT2i) | 180,740 vs 180,740 after 1:1 PSM | TriNetX | First prescription of GLP-1RA or SGLT2i | ICD-coded amputation (major, minor); DFU; mortality | Patient | 1:1 PSM; Kaplan-Meier; Cox | Favorable for GLP-1RA over SGLT2i. ↓ major LEA (HR 0.77), ↓ DFU (HR 0.92), ↓ mortality (HR 0.66). |
| *Selected comparator studies illustrating contrasts with the present analysis. CTS: carpal tunnel syndrome; DFU: diabetic foot ulcer; EHR: electronic health record; FDR: false discovery rate; GEE: generalized estimating equations; GLP-1RA: glucagon-like peptide-1 receptor agonist; HR: hazard ratio; LEA: lower-extremity amputation; NNT: number needed to treat; OR: odds ratio; PSM: propensity score matching; RR: relative risk; SGLT2i: sodium-glucose cotransporter-2 inhibitor; SSI: surgical site infection; T1DM: type 1 diabetes mellitus; T2DM: type 2 diabetes mellitus.* | | | | | | | | | |

**Table S4. Methodological comparison with representative GLP-1RA surgical outcomes studies.**

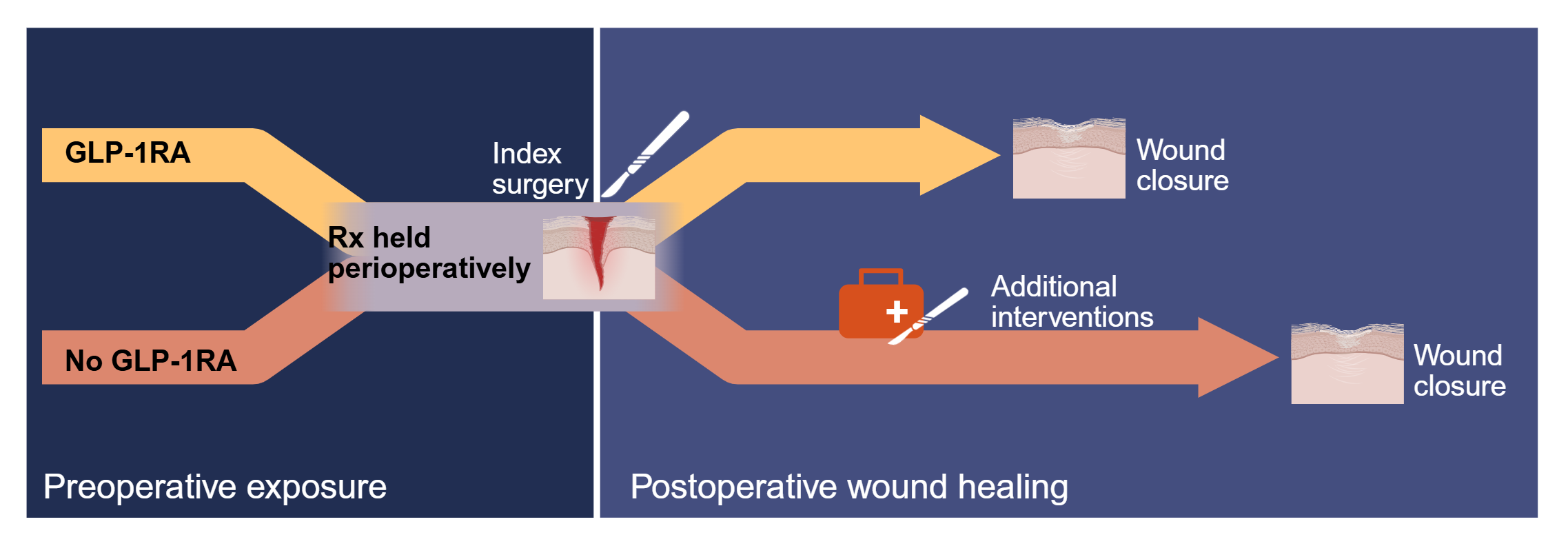
**Figure S1. Emerging model of preoperative GLP-1RA exposure and wound healing outcomes in the surgical course**. The timeline illustrates GLP-1RA preoperative exposure status, the perioperative window and associated postoperative wound healing outcomes indicated by this pilot study. GLP-1RA is typically paused in the perioperative period due to risks including aspiration and hypo- and hyperglycemia in the acute surgical window. This schematic reflects hypothesized biological relationships and does not represent causal claims.
